# Meplazumab treats COVID-19 pneumonia: an open-labelled, concurrent controlled add-on clinical trial

**DOI:** 10.1101/2020.03.21.20040691

**Authors:** Huijie Bian, Zhao-Hui Zheng, Ding Wei, Zheng Zhang, Wen-Zhen Kang, Chun-Qiu Hao, Ke Dong, Wen Kang, Jie-Lai Xia, Jin-Lin Miao, Rong-Hua Xie, Bin Wang, Xiu-Xuan Sun, Xiang-Min Yang, Peng Lin, Jie-Jie Geng, Ke Wang, Hong-Yong Cui, Kui Zhang, Xiao-Chun Chen, Hao Tang, Hong Du, Na Yao, Shuang-Shuang Liu, Lin-Na Liu, Zhe Zhang, Zhao-Wei Gao, Gang Nan, Qing-Yi Wang, Jian-Qi Lian, Zhi-Nan Chen, Ping Zhu

**Affiliations:** National Translational Science Center for Molecular Medicine & Department of Cell Biology, Fourth Military Medical University, Xi’an 710032, China; Department of Clinical Immunology, Xijing Hospital, Fourth Military Medical University, Xi’an 710032, China; Center for Infectious Diseases, Tangdu Hospital, Fourth Military Medical University, Xi’an 710038, China; Department of Clinical Diagnosis, Tangdu Hospital, Fourth Military Medical University, Xi’an 710038, China; College of Military Preventive Medicine, Fourth Military Medical University, Xi’an 710032, China; Jiangsu Pacific Meinuoke Biopharmceuticals Co. Ltd. Changzhou 213022, China; Department of Pharmaceutics, Tangdu Hospital, Fourth Military Medical University, Xi’an 710038, China; Department of Foreign Languages, Fourth Military Medical University, Xi’an 710032, China

## Abstract

**Background:** SARS-CoV-2 is a novel human coronavirus, there is no specific antiviral drugs. It has been proved that host-cell-expressed CD147 could bind spike protein of SARS-CoV-2 and involve in host cell invasion. Antibody against CD147 could block the infection of SARS-CoV-2. We aimed to assess the efficacy and safety of meplazumab, a humanized anti-CD147 antibody, as add-on therapy in patients with COVID-19 pneumonia.

**Methods:** All patients received recommended strategy from *Chinese Clinical Guidance for COVID-19 Pneumonia Diagnosis and Treatments* released by National Health Commission of China. Eligible patients were add-on administered 10 mg meplazumab intravenously at days 1, 2, and 5. Patients hospitalized in the same period were observed as concurrent control. The endpoints include virological clearance rate, case severity, chest radiographic, and laboratory test. This trial was approved by the Ethics Committee of Institution at the Tangdu hospital, and registered with ClinicalTrials.gov, NCT 04275245.

**Findings:** 17 patients were enrolled and assigned to meplazumab group between Feb 3, 2020 and Feb 10, 2020. 11 hospitalized patients served as concurrent control. Baseline characteristics were generally balanced across two groups. Compared to control group, meplazumab treatment significantly improved the discharged (p=0.005) and case severity (p=0.021) in patients. The time to virus negative in meplazumab group was reduced than that in control group (median 3, 95%CI[1.5-4.5] vs. 13, [6.5-19.5]; p=0.045, HR=0.374, 95%CI[0.143-0.978]). The percentages of patients recovered to the normal lymphocyte count and CRP concentration were also increased remarkably and rapidly in meplazumab group. No adverse effect was found in meplazumab-treated patients.

**Interpretation:** Meplazumab efficiently improved the recovery of patients with SARS-CoV-2 pneumonia with a favorable safety profile. Our results support to carry out a large-scale investigation of meplazumab as a treatment for COVID-19 pneumonia.

**Funding:** National Science and Technology Major Project.

## Introduction

Coronavirus is an enveloped positive stranded RNA virus belonging to the family *Cornonaviridae*, order *Nidovirales*.^1^ Over the past 20 years, two human coronaviruses, SARS-CoV and MERS-CoV were reported to lead to severe and fatal lower respiratory tract infection.^2,3^ In Dec 2019, an outbreak of a respiratory syndrome, COVID-19, was detected in China which is caused by a novel coronavirus, named SARS-CoV-2^4^ The genome sequence of SARS-CoV-2 shows it belongs to betacoronavirus genus, and has extremely high homology with SARS-CoV at genome and proteome.^5,6^

Illness onset among rapidly increasing numbers of COVID-19 in China and globe indicates that SARS-CoV-2 is more contagious than both SARS-CoV and MERS-CoV. The infection of SARS-CoV-2 leads to acute viral exudative pneumonia, with multiple organ damages, especially in lung, presenting bilateral diffuse alveolar damage with cellular fibromyxoid exudates.^7^ 80% cases have mild symptoms (including non-pneumonia and mild pneumonia cases), while about 20% patients have developed severe pneumonia and acute respiratory distress syndrome, which attribute to death.^8^ As of Mar 12th, 2020, 125,048 cases and 4,613 death have been reported globally.^9^

The current standard treatment including oxygen therapy, antiviral treatment, fluid management, and antimicrobial therapy for secondary bacterial infections.^10^ Although several drugs, such as remdesivir and chloroquine phosphate, have been registered in ongoing clinical trials for COVID-19, no specific antiviral drugs against this novel coronavirus are approved so far. The latest report indicates the infection of SARS-CoV-2 increases the expression levels of multiple proinflammatory cytokines, including IFN-γ, TNF-α, MCP1, and IL-6 in the serum, which suggests the inflammation storm may involve in the progression of COVID-19.^4^ Therefore, inhibiting virus replication and limiting excessive inflammation are of equal importance in COVID-19 therapy.

Angiotensin-converting enzyme 2 (ACE2) is the only reported host cellular receptor for SARS-CoV-2 to our knowledge,^11^ which presents in various human tissues, including alveolar epithelial cells, endothelial cells of arteries and veins, smooth muscle cells, etc.^12^ Our current published data showed a direct interaction between spike protein of SARS-CoV-2 and CD147, a type I transmembrane glycoprotein expressed on epithelial cells. A humanized IgG_2_ monoclonal antibody against CD147, meplazumab, effectively inhibited SARS-CoV-2 replication and virus-induced cytopathic effect in Vero E6 cells, both in a dose-dependent manner. These data demonstrated a novel entry of virus infection mediated by CD147 receptor.^13^

CD147 is also presented in activated inflammatory cells and participates the regulation of cytokine secretion and leukocytes chemotaxis by binding with cyclophilin A (CyPA).^14,15^ CyPA is a proinflammatory cytokine that is up-regulated in response to virus infection.^16^ Our previous studies demonstrated that antibody targeting CD147 inhibited CyPA-induced T cell chemotaxis thus attenuating the local inflammation.^17^

Based on these evidences, we assess the clinical outcomes using meplazumab as add-on therapy in patients with COVID-19 pneumonia. We designed an open-label, concurrent controlled trial to evaluate whether meplazumab infusion improves the patients with COVID-19 pneumonia by inhibiting virus replication and suppressing inflammation. We hope our study will shed light on finding a new drug and strategy to control this novel coronavirus-induced pneumonia.

## Methods

### Study design and patients

We did a prospective, single center, open-labelled trial at Tangdu Hospital of Fourth Military Medical University in Xi’an, China. The study protocol and consent were approved by the Independent Ethics Committee of Institution for National Drug Clinical Trials at the Tangdu hospital (K202002-01). The study was registered at ClinicalTrials.gov (NCT04275245), before any patient enrollment.

Enrolled patients fulfilled inclusion and exclusion criteria. The inclusion criteria are as follows: men and women aged 18 to 78 years; patients with common, severe, or critical COVID-19 pneumonia were laboratory and clinical diagnosed according to *Chinese Clinical Guidance for COVID-19 Pneumonia Diagnosis and Treatments* released by National Health Commission of China;^18^ the subjects must understand the study and be willing to participate in the study. The exclusion criteria are as follows: allergic reactions or a history of allergy to any of the ingredients treated in this trial; patients not suitable to participate in this study by the judgment of the investigator. Each patient signed an informed consent form before enrollment.

Case severity of COVID-19 is categorized as common, severe, and critical by the vital signs, oxygenation index (PaO_2_/FiO_2_), chest radiographic, and vital organ function based on the *Diagnosis and Treatment for 2019 Novel Coronavirus Disease*.^18^ Common case was featured by fever, respiratory symptoms, and radiographic pneumonia. Severe case was characterized by any of the following signs: dyspnea, respiratory frequency ≥30/minute, blood oxygen saturation ≤93%, and PaO_2_/FiO_2_ ratio <300mmHg. Critical case was characterized by any of the following signs: respiratory failure needing mechanical ventilation, shock, and multiple organ dysfunction/failure needing Intensive Care Unit (ICU). The patients who met all the following criteria will be discharged: body temperature recovered and remained normal more than 3 days; respiratory symptoms relief, and two continuous negative for nasopharyngeal swab test for SARS-CoV-2 (interval more than 24 hours).

In same period, hospitalized patients in the same center were observed as concurrent control, and were required to follow the inclusion and exclusion criteria mentioned above.

### Procedure

Any enrolled patients received recommended treatment according to the *Diagnosis and Treatment for 2019 Novel Coronavirus Diseases*.^18^ Physicians were allowed to use any necessary treatment, laboratory, and radiographic examination with their standard of care. Clinical, laboratory, and radiographic assessments were conducted at baseline, including complete blood count, serum biochemical test (renal function, liver function, and C reactive protein (CRP)), serum electrolytes test, coagulation analysis (PT, APTT, INR, FIB, D-Dimer), chest CT/chest X-ray, nasopharyngeal swab test for SARS-CoV-2 (using qRT-PCR assay approved by the National Medical Products Administration).

There were 18 study visits from baseline to day 28. 10mg meplazumab was administered on day 1, day 2 and day 5 by intravenous infusion within 60-90 min. Efficacy and safety were assessed at baseline, every day after day 1 to day 14, and every week thereafter up to day 28 or discharge.

### Outcomes

The primary study endpoint was the virological clearance (i.e. negative conservation rate and time to negative) using qRT-PCR in nasopharyngeal swabs samples. Secondary efficacy endpoints were assessment of time to recovery of vital sign (including body temperature, respiratory rate, and SPO_2_), chest radiographic improvement, rate of PaO_2_/FiO_2_ recovery, time (days) to discharge, and inflammation recovery (percentage of patient with normal CRP concentration).

### Statistical analysis

Continuous variables were expressed as median (IQR) and compared with the Mann-Whitney U test. Categorical variables were expressed as number (%) and compared by Fisher’s exact test, Ordinal regression, or McNemar’s test. 100% stacked column were drawn to describe composition of case severity, chest radiographic improvement, lymphocyte count, and CRP concentration. Differences between discharged and virus nucleic acid negative conversion curves were analyzed by Cox regression analysis, and multiple cox regression was also used to take account of potential confounding variables including group, age, case severity, and glucocorticoid treatment. A two-sided α of less than 0.05 was considered statistically significant. Statistical analyses were performed using the SPSS software, version 23.0 and GraphPad Prism software, version 5.0.

## Results

Between Feb 3, 2020, and Feb 10, 2020, we screened 39 patients diagnosed with COVID-19 in Tangdu Hospital. The reasons for screen failure are listed in figure 1. 17 patients signed informed consent and were allocated to meplazumab group. 11 hospitalized patients who met the inclusion criteria and with no exclusion criteria signs were collected as concurrent control in the same period. Demographics, comorbidity, epidemiological exposure, prescribed treatment, signs & symptoms, and case severity were generally balanced across two groups as shown in table 1. The median age of patients was 51 years (IQR 49-67) in meplazumab group and was 64 years (IQR 43-67) in control group. Patients in both groups had epidemiological exposure history. Underlying comorbidity, including diabetes, hypertension, cardiovascular disease, and Parkinson disease, were observed in 52.9% and 36.4% patients in meplazumab and control groups, respectively. All patients had received recommended therapy according to the *Diagnosis and Treatment for 2019 Novel Coronavirus Diseases*,18 including antiviral treatment, glucocorticoid treatment, and antibiotic treatment. 4 common cases, 6 severe cases, and 7 critical cases were enrolled in meplazumab group. In contrast, 4 common cases, 4 severe cases, and 3 critical cases were collected in control group, showing no difference compared to meplazumab group (p=0.688). Subsequently, 11 patients had received 3 doses meplazumab, and 6 patients had received 2 doses meplazumab.

**Table 1:** Demographics and baseline characteristics of patients infected with SARS-CoV-2

|  | **Meplazumab**<br>**(n = 17)** | **Control**<br>**(n = 11)** | **p**<br>**value** |
| --- | --- | --- | --- |
| **Characteristics** |  |  |  |
| Age, years | 51 (49-67) | 64 (43-67) | 0.981 |
| Sex |  |  | 0.441 |
| Women | 6 (35.3%) | 6 (54.5%) |  |
| Men | 11 (64.7%) | 5 (45.5%) |  |
| Current smoking | 1 (5.9%) | 1 (9.1%) | 1.000 |
| Epidemiological exposure |  |  | 0.934 |
| No exposure | 6 (35.3%) | 5 (45.5%) |  |
| Hubei exposure | 10 (58.8%) | 4 (36.4%) |  |
| Confirmed patient exposure | 1 (5.9%) | 2 (18.2%) |  |
| Any comorbidity | 9 (52.9%) | 4 (36.4%) | 0.460 |
| Diabetes | 3 (17.6%) | 0 (0%) | 0.258 |
| Hypertension | 6 (35.3%) | 3 (27.3%) | 1.000 |
| Cardiovascular disease | 1 (5.9%) | 2 (18.2%) | 0.543 |
| Chronic obstructive pulmonary disease | 1 (5.9%) | 0 (0%) | 1.000 |
| Parkinson disease | 1 (5.9%) | 0 (0%) | 1.000 |
| **Treatment** |  |  |  |
| Antiviral treatment |  |  |  |
| Lopinavir and Ritonavir | 17 (100%) | 11 (100%) | NA |
| Recombinant human interferon α-2b | 17 (100%) | 11 (100%) | NA |
| Glucocorticoid | 16 (94.1%) | 7 (63.6%) | 0.062 |
| Antibiotic | 17 (100%) | 10 (90.9%) | 0.393 |
| **Signs and symptoms** |  |  |  |
| Fever | 16 (94.1%) | 11 (100%) | 1.000 |
| Highest temperature, °C |  |  | 0.914 |
| <37.3 | 1 (5.9%) | 0 (0%) |  |
| 37.3-38.0 | 9 (52.9%) | 7 (63.6%) |  |
| 38.1-39.0 | 5 (29.4%) | 2 (18.2%) |  |
| >39.0 | 2 (11.8%) | 2 (18.2%) |  |
| Cough | 9 (52.9%) | 7 (63.6%) | 0.705 |
| Myalgia or fatigue | 10 (58.8%) | 5 (45.5%) | 0.700 |
| Sputum production | 1 (5.9%) | 2 (18.2%) | 0.543 |
| Diarrhoea | 2 (11.8%) | 1 (9.1%) | 1.000 |
| Dyspnoea | 10 (58.8%) | 7 (63.6%) | 1.000 |
| Respiratory rate >24 breaths per min | 7 (41.2%) | 5 (45.5%) | 1.000 |
| **Case severity** |  |  | 0.388 |
| Common | 4 (23.5%) | 4 (36.4%) |  |
| Severe | 6 (35.3%) | 4 (36.4%) |  |
| Critical | 7 (41.2%) | 3 (27.3%) |  |

Case severity is categorized according to the Chinese Clinical Guidance for COVID-19 Pneumonia Diagnosis and Treatments released by the National Health Commission of China. Data are median (IQR) or n (%). p values comparing meplazumab and control are from Fisher's exact test, Mann-Whitney U test, or Ordinal regression. NA = not applicable.

**Figure 1:**
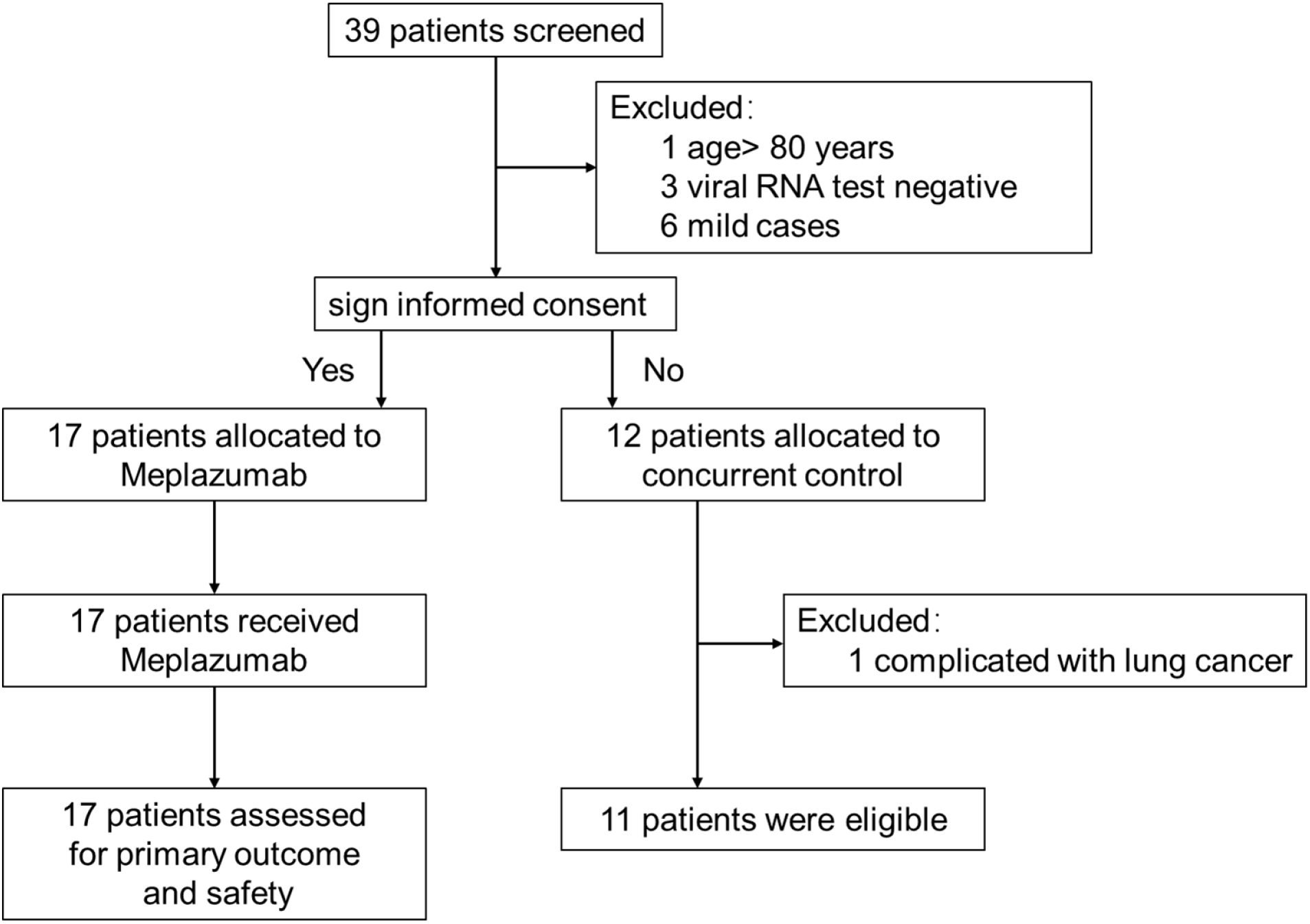
Trial profile.

The distribution of case severity and discharged cases was analyzed in each group over 28 days follow-up as shown in table 2. The overall improvement rates (defined as the percentage of discharged or improved cases) at days 7, 14, 21, 28 were 17.6% (3/17), 47.1% (8/17), 82.4% (14/17), and 94.1% (16/17) in the meplazumab group, while those were 0% (0/11), 27.3% (3/11), 54.5% (6/11), and 81.8% (9/11) in the control group, respectively. One aggravating case was observed in control group at day 7. At the end of observation (day 28), both groups showed significant improvement compared to baseline (p<0.001 for both meplazumab and control).

**Table 2:**
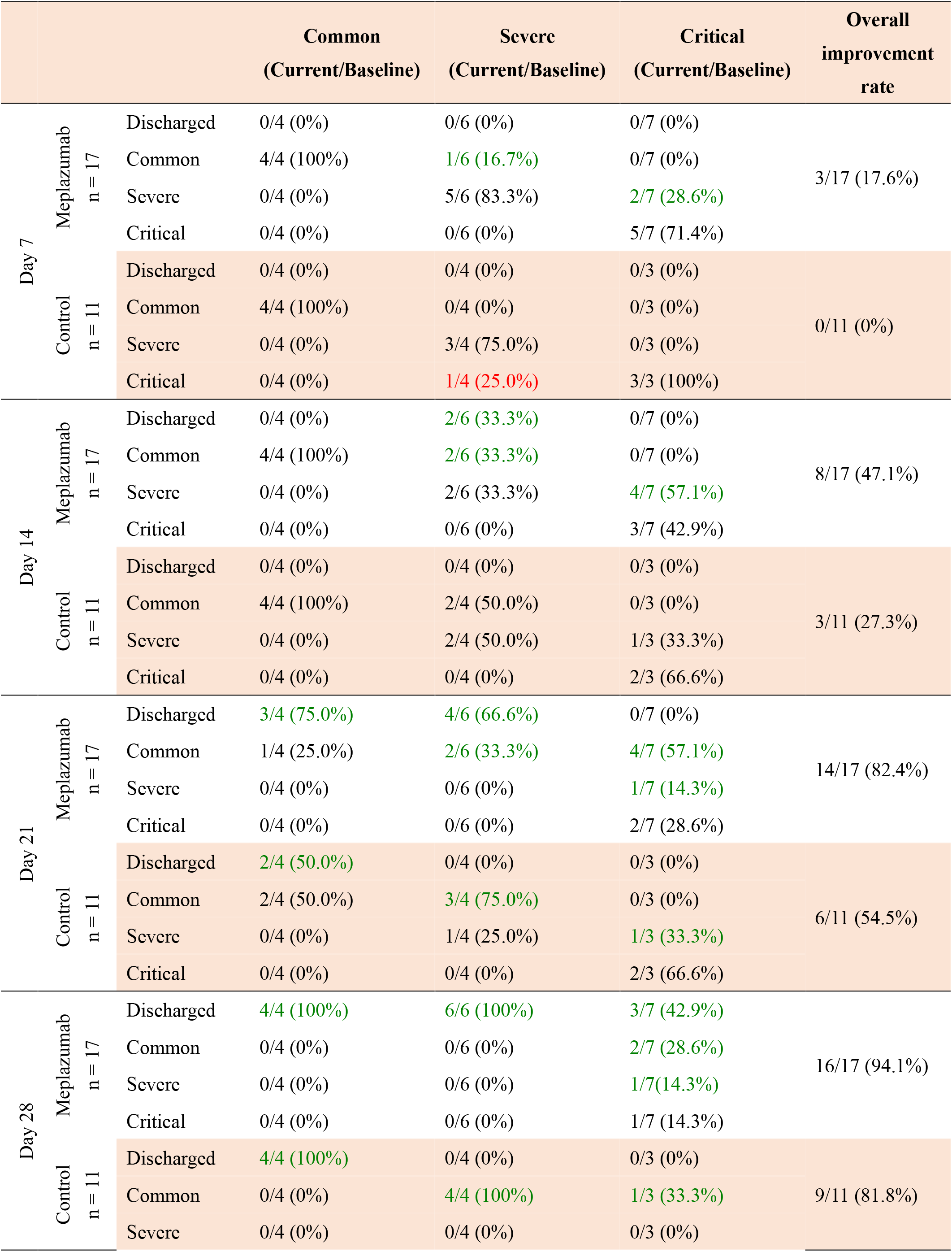
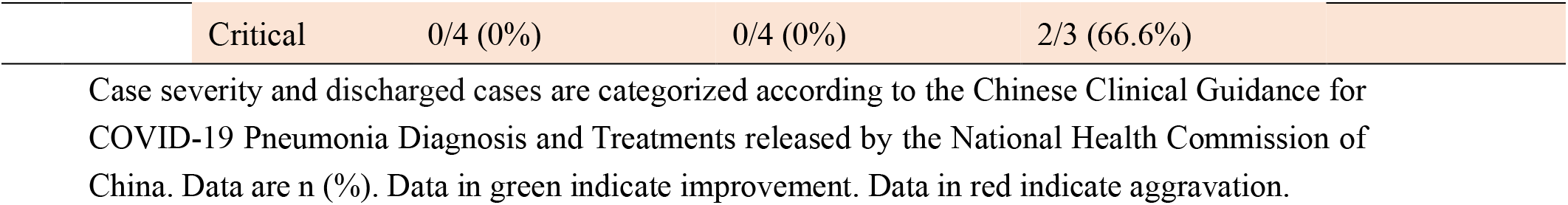
Distribution of case severity and discharged cases

To observe the efficacy of meplazumab add-on therapy, we analyzed the time to discharge using multiple Cox regression analysis. As shown in figure 2A, the meplazumab treatment significantly improved the discharged of cases (p = 0.005) compare to control cohort.

**Figure 2:**
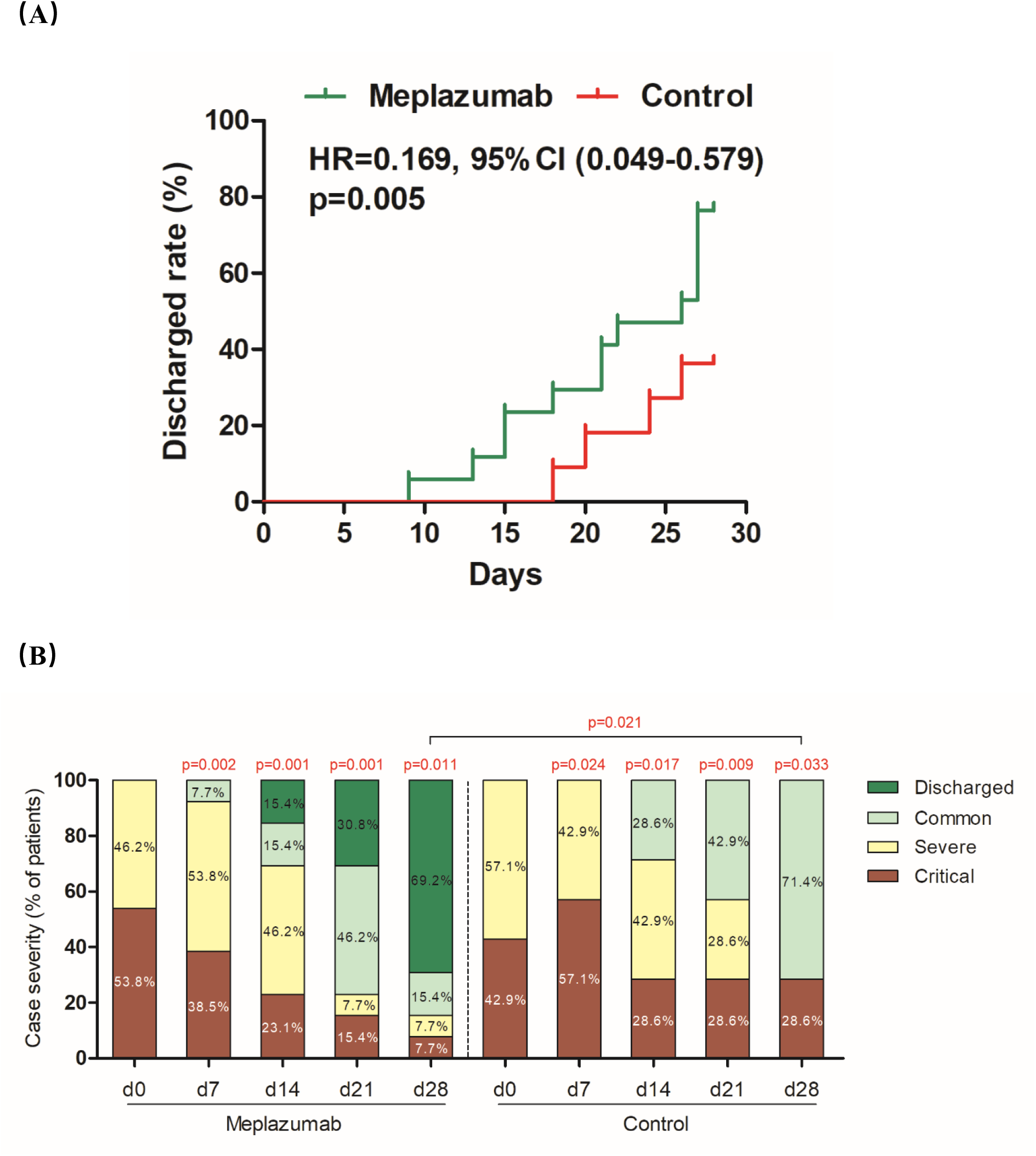
Case discharge and case severity in patients. (A) Analysis of time-to-discharge in patients. The p value was determined using Cox regression analysis. (B) Distribution of case severity in severe and critical patients. The p values were from Ordinal regression. The p value comparing between baseline (d0) and each time point was shown on the top of column, and p value comparing between meplazumab and control at day 28 was shown on the line.

Recovery in severe and critical cases is of paramount importance to reduce mortality of COVID-19. The case severity of severe and critical cases in both groups were improved significantly compared to baseline, since 7 days post-treatment (figure 2B). At day 28, 4 severe cases and 1 critical case were improved to common and no case was discharged in control group. In meplazumab group, 9 cases (6 severe and 3 critical) were discharged, 2 critical cases were improved to common, and 1 critical case was improved to severe, demonstrating a significantly beneficial outcome compared to control group (p=0.021, table 2 and figure 2B). These results indicated that meplazumab treatment accelerated the improvement and make a rapid recovery from COVID-19 pneumonia, especially for the severe and critical cases. In this trial, no death was reported in all patients.

Chest radiographic analysis was performed independently by two radiologists, and graded by the changed areas of ground-glass opacity and consolidation compared with the baseline. As shown in figure 3, the chest radiographic, compared to baseline in both two groups, were improved gradually over 28 days. At day 7, in the meplazumab group, 1 case (5.9%, 1/17) improved more than 50%, and 7 cases (41.2%, 7/17) improved 25-50%, while no patients improved over 25% in control group. At day 14, 47.1% patients (8/17) in meplazumab group improved more than 50%, and no similar situation was observed in control group. At day 21, 12 case (70.6%, 12/17) improved more than 50%, and 3 cases (17.6%, 3/17) improved 25-50% in meplazumab group, while 3 case (27.3%, 3/11) improved more than 50%, and 5 cases (45.5%, 5/11) improved 25-50% in control group. The meplazumab group showed more significant benefit than control group in days 7, 14, and 21 (p=0.010, p=0.006, and p=0.037, respectively), which coincident with the improvement of case severity. A representative chest CT image of meplazumab-treated patient was shown in figure 4, which displayed the CT imaging manifestations (bilateral ground-glass shadow and consolidation) of COVID-19 at baseline period, and all these lesions were resolved at 22 days post-treatment.

**Figure 3:**
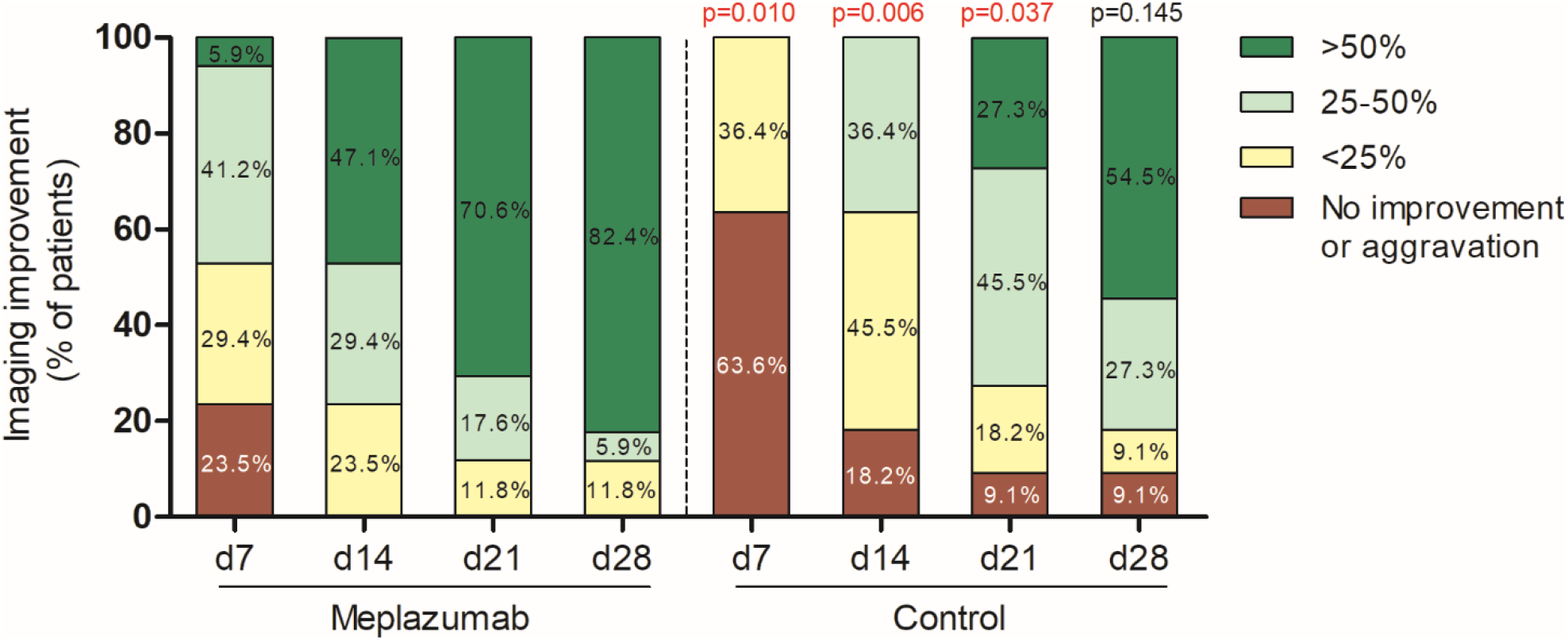
Chest imaging analysis. The p values comparing between meplazumab and control at days 7, 14, 21, and 28 were from Ordinal regression.

In this trial, the clearance of virus has been evaluated by negative-conversion rate and the time to negative. At day 7, the rate of virus nucleic acid negative conversion in control group was 27.3% (3/11), and reached to 54.4% (6/11) at day 14. While, the rate was 76.5% (13/17) in meplazumab group at day 7, and reached to 94.1% (16/17) at day 14, which were significantly higher than control group (p=0.019 and p=0.022, respectively). The analysis of time to virus negative indicated that meplazumab-treated patients converted to negative in a shorter period than patients in control group significantly (median 3, 95%CI[1.5-4.5] vs. 13, [6.5-19.5]; p=0.045, HR=0.374, 95%CI[0.143-0.978], figure 5). The results of multiple Cox regression indicate that no baseline characteristics, including age (p = 0.092), glucocorticoid treatment (p = 0.339), and case severity (p = 0.455) contributed to the difference of time to virus negative. All these data indicate an obvious benefit of meplazumab treatment in the clearance of SARS-CoV-2.

**Figure 4:**
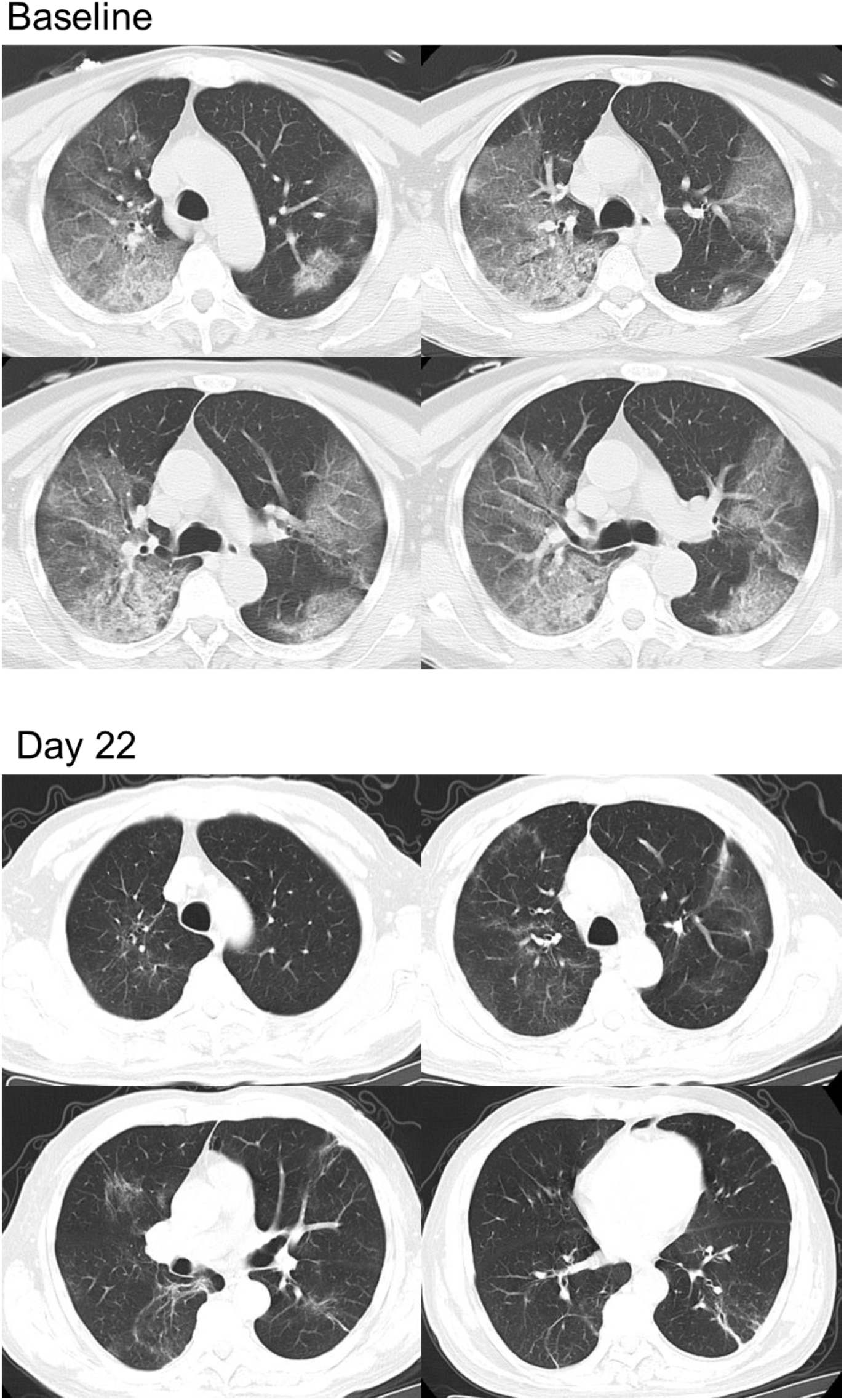
Chest CT image. Transverse chest CT images from a 65-year-old male patient showing bilateral large flaky ground glass shadows and consolidation on baseline period. At day 22, the intrapulmonary lesions were significantly absorbed and dissipated by meplazumab add-on treatment.

**Figure 5:**
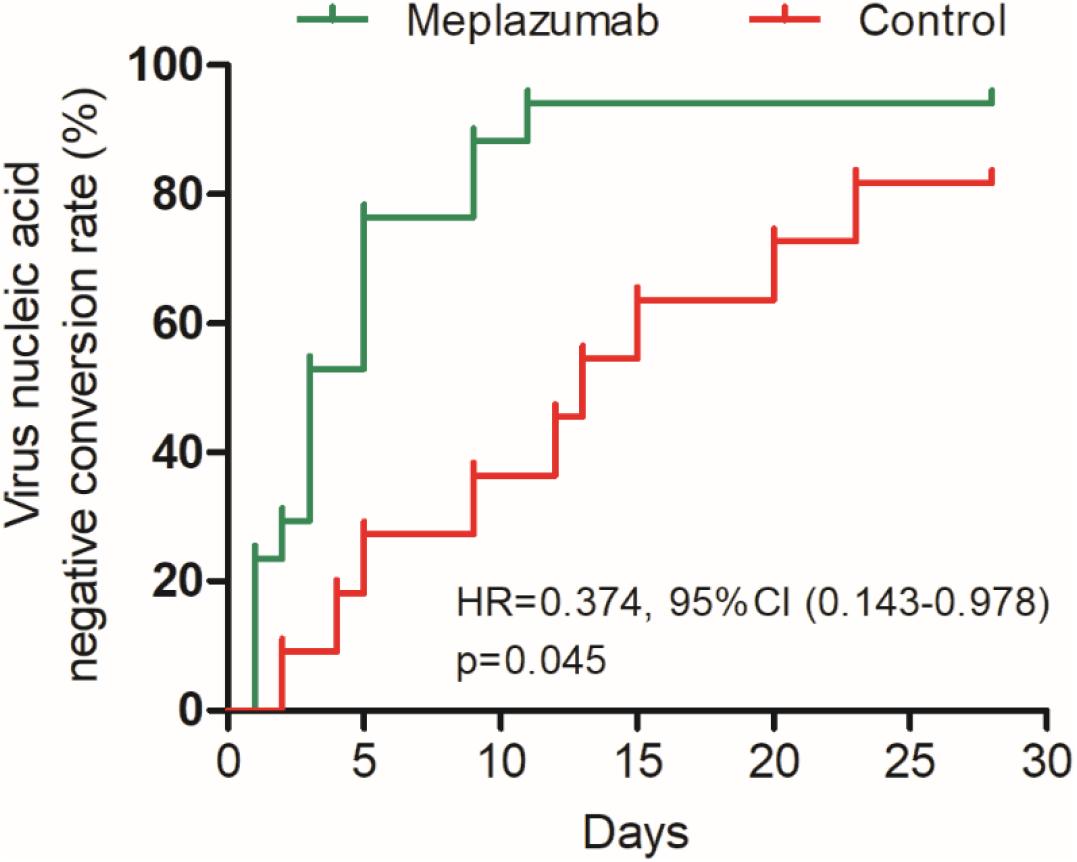
Time to virus negative conversion. Analysis was performed by Cox regression analysis, with p value 0.045 and HR 0.374, 95%CI[0.143-0.978]). Median of time-to-negative was 3 days (95%CI[1.5-4.5]) for meplazumab group, and 13 days (95%CI [6.5-19.5]) for control group.

It was reported that lymphocytopenia is a typical characteristics and correlated with the prognosis of COVID-19 pneumonia. In our study, we compared the lymphocyte count between baseline and post-treatment in both groups. As shown in figure 6A, in observation period (day 7 to 28), the percentages of patients with a normal lymphocyte count (>0.8×10^9^/L) were increased in both groups, while the improvement in meplazumab group was more notable. Compared to baseline, the percentage in meplazumab group was improved significantly as early as day 7 (p=0.031), but no significance difference was detected in control group over the follow-up period.

**Figure 6:**
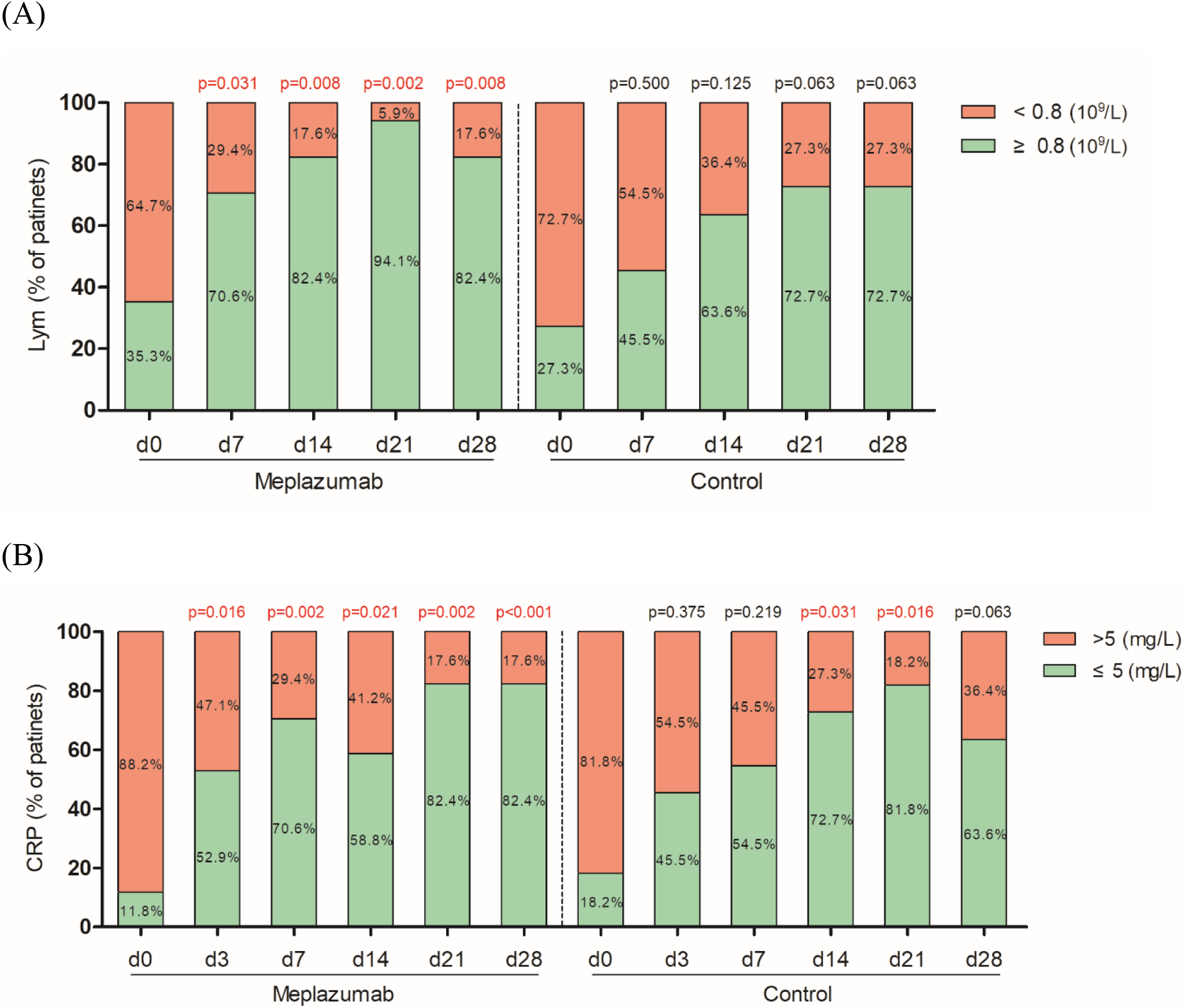
Proportion of patients regarding to lymphocyte count and CRP concentration. Proportion of patients (% patients) with or without normal lymphocyte count (A) and CRP concentration (B). The p value comparing between baseline (d0) and each time point was determined by McNemar’s test, shown on the top of column.

We also measured the concentration of CRP, as a predictors of COVID-19 severity.^19^ As shown in figure 6B, from day 3 to day 28, the percentages of patients with a normal CRP concentration (≤5mg/L) were increased from 52.9% (9/17) to 82.4% (14/17), indicating significant increases compared to baseline (all p<0.05 at days 3, 7, 14, 21, and 28). In control group, the significant increases were observed at day 14, and day 21 compared to baseline. The data suggests that meplazumab exhibited an effect on the control of virus-induced acute inflammation at early management.

In meplazumab group, the elevation of both ALT and AST (≥ 2 ULN) was reported in 2 patients (2/17, 11.8%). In control group, both ALT and AST elevations were reported in 1 case (9.1%), and single ALT elevation was reported in 1 case (9.1%). The increased level of ALT and AST returned to normal after 7 days in all four patients, and the treatment procedure was not affected by their fluctuation. The abnormal transaminase was not related with meplazumab treatment after comprehensive judgement by the investigators. No other adverse event was reported in meplazumab-treated patients, including reaction around the injection site, rash, nausea, vomit, anemia, neutropenia, thrombocytopenia, total bilirubin, albumin and creatinine etc.

## Discussion

With the rapid escalation of the COVID-19, over 120 countries have reported more than 140,000 cases in the world. As of Mar 8, 2020, 73 clinical trials have been registered in Clinicaltrial.gov, in which nearly one fifth reagents are small molecular compounds, such as remdesivir and oseltamivir, and four are antibody drugs like PD-1 blocking antibody, bevacizumab, eculizumab, and meplazumab.^20^ Here, we reported an add-on trial, using anti-CD147 humanized monoclonal antibody, meplazumab, with a novel pharmacological mechanism to treat COVID-19 pneumonia by blocking virus invasion and attenuating inflammation. The adding meplazumab 20-30 mg in patients with COVID-19 who have received recommended treatment,^18^ increased the virological clearance rate, promoted the recovery of chest radiographic and lymphocytopenia, decreased the inflammation index (CRP), accelerating the improvement of disease without serious adverse event.

In our recent published study, we have revealed that CD147 is a novel identified receptor for SARS-CoV-2 infection by binding with spike protein. SARS-CoV-2 infection was efficiently inhibited by meplazumab targeting CD147 in a dose dependent manner. All these results provide insights that CD147 acts as a functional entry receptor for SARS-CoV-2, and serves as a therapeutic target for inhibiting virus infection.^13^

It was reported 18.5% cases in COVID-19 were categorized as severe and critical cases, and fatality rate for critical case was 49.0% to 61.5%.^4,8^ These studies reveal the great challenge of enhancing the therapeutic effects for severe case of COVID-19, and focus on how to conduct effective intervention as early as possible. In this add-on trial, we introduced a novel therapy target to block the virus infection and inflammation, with a favorable efficacy. Meplazumab treatment accelerated the improvement of COVID-19, especially in the severe and critical cases. At the day 28, 69.2% severe and critical cases were discharged, and no death case was reported. The improvement of chest imaging, clearance of virus, and recovering of lymphocyte count, were observed within 1 week management, which indicated that the therapeutic effects facilitated prognosis.

CD147 is a receptor for ligand CyPA, and its interaction with CD147 was key to the inflammation and chemotaxis.^21^ CyPA is secreted to extracellular in response to inflammatory stimuli (e.g. virus infection), activates and attracts leukocytes via its receptor CD147 to the stimulus site.^15^ We demonstrated that anti-CD147 antibody could attenuate the chemotactic index of T cells induced by CyPA.^17^ Multiple cytokines and chemokines increased significantly in patients with COVID-19, which was significantly correlated with pulmonary inflammation index of chest CT imaging.^4,22^ These results suggest that the excess immune cells migrating into lung tissue may cause uncontrolled immune response, leading to the inflammation storm, and aggravated disease. We presumed that meplazumab blocked the interaction between CyPA and CD147, attenuated the chemotactic effect of CyPA, decreased the immune cells in lung tissue, and facilitated the improvement of chest radiographic. In this trial, the remarkable and rapid resolution of inflammatory foci in lungs, like ground-glass opacity and consolidation, were observed in meplazumab treatment group.

Lymphopenia was common in patients with COVID-19 and SARS patients, and can be used as an indicator of disease severity and prognosis.^23^ 75.4% patients with COVID-19 reported lymphopenia.^24^ Due to the absence of ACE2 in lymphocyte, it has been proposed that the lymphocytopenia in SARS patient is caused by the indirect mechanism, including inflammation storm, vascular cell adhesion, soluble Fas ligand, and glucocorticoids.^12,25^ However, the SARS-CoV particles and genomic sequence were detected in a large number of circulating lymphocytes, causing destruction of lymphocytes.^26^ In this trial, lymphocyte count in meplazumab-treated patients were restored in short time. CD147 is high expressed on the activated T cells,^17^ which may facilitate the invasion of SARS-CoV-2 to lymphocytes by binding spike protein, suggesting that CD147 may be involved in lymphocytopenia. Meplazumab interrupted this process by preventing virus invasion to keep lymphocytes survived. Second, the meplazumab blocking CD147-CyPA interaction might contribute to lymphocyte elevation in peripheral blood by inhibiting the lymphocyte accumulation in pulmonary organ.

In summary, we establish a “specific antibody add-on therapy model”, a combined therapeutic strategy of the specific antibody with recommended antivirus treatment, and demonstrate it safe and effective. The effective dosage of antibody by timely administration attenuates the inflammation in patients with COVID-19, and facilitates the patients to get through acute exudation of pneumonia. Therefore, antibody therapy can extend the therapeutic window, thus safeguard patients pass the dangerous period.

A favorable specific antibody drug should inhibit the virus replication and suppress the inflammation storm. Meplazumab, an anti-CD147 antibody, can block the receptor of host cells, and inhibit the virus invasion and replication by interrupting spike protein from recognizing CD147 receptor. Meanwhile, meplazumab blocks the CD147, the receptor of proinflammatory factor CyPA, thus suppressing the inflammation.

Although we get the outcomes of meplazumab treatment for COVID-19, our results are limited by the small subject size and the absence of parallel control. Therefore, confirmation from larger scale and randomized, double-blind, placebo-controlled trial is required to fully assess the efficacy and safety of meplazumab in patients with COVID-19 pneumonia.

## Data Availability

The data that support the findings of this study are available from the corresponding author, PZ, upon reasonable request.

## Contributors

PZ, ZNC, and HB were responsible for initial study design. JQL, ZHZ, DW, ZZ, CQH, WZK, WK, HD, NY, and LNL were responsible for study implementation and enrolment of participants. KD, ZZ, and ZWG were responsible for laboratory tests. RHX, BW, XXS, XMY, PL, JJG, KW, HYC, KZ, XCC, HT, and SSL were responsible for data collection and laboratory technique support. JLX, JLM, and GN contributed to data analysis. QYW contributed to English writing. PZ, ZNC, HB, ZHZ, and DW analyzed and interpreted the data, wrote and revised the manuscript. All authors had opportunity to review the data and edited the final report.

## Acknowledgments

The authors thank the study participants, physicians, nurses, clinical research officers, and laboratory technicians that were provided throughout the conduct of this study. This study was funded by the National Science and Technology Major Project (2019ZX09732-001).

